# Widespread Cortical Thickness Reductions Following Non-medical Use of Ketamine: a Structural MRI Study of Individuals with Ketamine Dependence

**DOI:** 10.1101/2021.02.21.21252178

**Authors:** Jinsong Tang, Qiuxia Wu, Chang Qi, An Xie, Jianbin Liu, Yunkai Sun, Tifei Yuan, Wei Chen, Tieqiao Liu, Wei Hao, Yanhui Liao

## Abstract

**Background:** A version of ketamine, called Esketamine has been approved for treatment-resistant depression (TRD). Ketamine (“K powder”), a “dissociative” anesthetic agent, however, has been used non-medically alone or with other illicit substances. Our previous studies showed a link between non-medical ketamine use and brain structural and functional alterations. We found dorsal prefrontal gray matter reduction in chronic ketamine users. It is unknown, however, whether these observations might parallel findings of cortical thickness alterations. This study aimed at exploring cortical thickness abnormalities following non-medical, long-term use of ketamine.

**Methods:** Structural brain images were acquired for 95 patients with ketamine dependence, and 169 drug-free healthy controls. FreeSurfer software was used to measure cortical thickness for 68 brain regions. Cortical thickness was compared between the two groups using analysis of covariance (ANCOVA) with covariates of age, gender, educational level, smoking, drinking, and whole brain mean cortical thickness. Results were considered significant if the Bonferroni corrected P-value < 0.01.

**Results:** Compared to healthy controls, patients with ketamine dependence have widespread decreased cortical thickness, with the most extensive reductions in the frontal (including the dorsolateral prefrontal cortex, DLPFC) and parietal (including the precuneus) lobes. Increased cortical thickness was not observed in ketamine users relative to comparison subjects. Estimated total lifetime ketamine consumption is correlated with the right inferior parietal and the right rostral middle frontal cortical thickness reductions.

**Conclusions:** This study provides first evidence that, compared with healthy controls, chronic ketamine users had cortical thickness reductions.

## Introduction

Ketamine is a water-soluble Phencyclidine or PCP [1-(1-phenylcyclohexyl)-piperidine] derivative, which functions mainly as an antagonist of the N-methyl-d-aspartate (NMDA) receptor. The ketamine molecule contains an asymmetric carbon atom with two enantiomers: the S (+) isomer and the R (−) isomer (1, 2). In general, the S (+) isomer ketamine is believed to be more potent than the R (−) isomer ketamine. The S-ketamine has an approximately four times greater affinity for the NMDA receptor (2), more potential benefit of anesthetic and analgesic (1), and of regaining consciousness and orientation (3) than the R-ketamine. The R-ketamine may be primarily associated with negative emergence reactions, such as alteration of arousal and breathing, disorientation and hallucinations (4).

Ketamine has a wide variety of clinical applications, such as its well-established use for anesthesia (approved by the U.S. Food and Drug Administration (FDA) in 1970 for human anesthesia, in particular to provide anesthesia in out-of-hospital emergencies, disaster situations and low-resource countries), pain management, and critical care (5-7). In 2000, a placebo-controlled, double-blinded trial reported the potential treatment effects of a single dose of ketamine in patients with depression (8). Since then, a series of preclinical and clinical studies have demonstrated ketamine’s role for the treatment of depression. During the past several years, a growing number of trials have demonstrated single and repeated ketamine infusions produce rapid and robust transient antidepressant for the treatment of severe depression (9-11). In March 2019 the FDA approved a new nasal spray medication (Esketamine/S-ketamine) for treating depression, but because of safety concerns (such as dizziness, euphoria, sedation, dissociation, misuse potential and addictive potential), it is used only for treatment-resistant depression (TRD) and in conjunction with an oral antidepressant, and available only at a certified doctor’s office or clinic (12-15).

Ketamine, as an addictive substance, has been non-medically used (by snorting the powder, called “K powder”) alone or with other illicit substances in both China and many other parts of the world (16). The common mental and physical adverse effects of non-medical use of ketamine (as a “club drug”) including psychotic symptoms and cognitive impairment (17), out-of-body experiences (18), depressive symptoms and anxiety symptoms (19), sleeping disturbances (20), ulcerative cystitis (21), and gastrointestinal toxicity (22). Administration of ketamine exacerbated psychotic symptoms and cognitive impairment in patients with schizophrenia (23-25), and induced hallucinations (25, 26) or perceptual distortions (27) in healthy people. Ketamine provided a unique model of psychosis or schizophrenia in animals (28).

Our previous studies demonstrated that chronic, non-medical use of ketamine, especially longer-term non-medical use and larger amounts of ketamine consumption, has been associated with dorsal prefrontal gray matter reduction (29), frontal white matter abnormalities (30), decreased thalamocortical connectivity (31), and regional homogeneity (ReHo) alterations of resting-state brain activity (32). It is unknown, however, whether these observations might parallel findings of cortical thickness alterations to the prefrontal, and whether brain structural alternation would be associated with ketamine use variables, such as age of ketamine use onset, the duration of ketamine use, and the quantity of ketamine consumption.

As the treatment with ketamine has not yet undergone the test of large-scale controlled trials to demonstrate the durability and safety of long-term treatment, there is limited evidence that long-term use of ketamine can be approved to individuals suffering from serious depression, or treatment can provide significant long-term benefits to this group of patients. Measuring the consequences of long-term, non-medical use of ketamine on brain changes may provide some insight for its long-term use as an antidepressant. This study aimed at exploring non-medical use of ketamine-induced cortical thickness alterations in patients with ketamine dependence/addiction. Considering non-medical ketamine use has been associated with poor psychological well-being (such as cognitive impairment, depression and anxiety) (33-35), acute (36) and chronic (37) use of ketamine mimic some aspects of schizophrenia, and widespread reductions of cortical thickness were observed in patients with schizophrenia (38), especially prefrontal cortical thickness abnormalities link to negative symptoms in schizophrenia (39), we hypothesized that, compared with healthy controls, ketamine dependent patients exhibit widespread cortical thickness reductions across the brain with prefrontal regions being generally more affected than others areas; regional cortical thickness alternations are associated with clinical symptoms (such as larger amount of ketamine consumption, longer-term use) in patients with ketamine dependence.

## Methods

### Study samples

The current study included a total of 95 patients with ketamine dependence from two drug rehabilitation centers in China (the Kangda Voluntary Drug Rehabilitation Centre in Hunan Province and the Department of Addiction Medicine, Hunan Brain Hospital) and 169 age-matched healthy volunteers via a combination of targeted site sampling, social media advertisement and snowball sampling referrals. Subjects were Chinese speaking, aged 18-45 years old. Ketamine dependent subjects and drug-free healthy control subjects were excluded if they: 1) had learning disabilities or central nervous system dysfunctions; 2) had any current or previous major medical or psychiatric disorders (excluding ketamine use induced psychiatric symptoms); 3) current use intravenous drugs; 4) had undergone current or previous use of electroconvulsive therapy (ECT) or brain stimulation therapies; 5) had a history of head injury with skull fracture or a loss of consciousness for more than 10 minutes; 6) had a family history of psychotic disorder; 7) met substance dependence diagnosis (excluding ketamine and nicotine for subjects in the ketamine use group, nicotine for subjects in the control group); 8) pregnancy or contraindications for MRI (Magnetic resonance imaging).

Ketamine dependent subjects used ketamine for at least one year. Lifetime ketamine dependence diagnosis was based on the Diagnostic and Statistical Manual of Mental Disorders (DSM-IV) criteria using the Structured Clinical Interview for DSM Disorders (SCID) (40), by trained clinicians (two licensed psychiatrists, at MD level). All individuals completed demographic and clinical information, and structural imaging data.

Each study sample was collected with subjects’ written informed consent approved by the Ethics Committee of the Second Xiangya Hospital, Central South University (No. S163, 2011). The authors assert that all procedures contributing to this work were carried out in accordance with the Helsinki Declaration of 1975, as revised in 2008.

### Clinical measurements

The sociodemographic characteristics were collected including age, gender, educational level, ethnicity, handedness, and marital status. The addictive substance use characteristics were collected including ketamine use, cigarette smoking, alcohol drinking and other illicit drug use variables. We applied a structured interview guide for the 30-item Positive and Negative Syndrome Scale (PANSS) (41), the 24-item Hamilton Depression Rating Scale (HDRS) (42, 43) and the 14-item Hamilton Anxiety Rating Scale (HARS) (44-46). All psychiatrists (Dr. Liao, Dr. Tang, Dr. Qi and Dr. Wu) who participated in the evaluation received training in the application of PANSS, HDRS and HARS before the study.

### MRI data acquisition

Three-dimensional T1-weighted images (3D-T1WIs) were obtained using a 3.0-T Siemens Magentom Trio scanner (Allegra; Siemens, Erlangen, Germany) located at the Magnetic Resonance Center of Hunan provincial People’s Hospital, China. The MPRAGE (magnetization-prepared rapid acquisition with gradient echo) sequence parameters of the structure T1-weighted images were: 176 sagittal slices of 1 mm thickness without gap, repetition time = 2000 ms, echo time = 2.26 ms, field of view = 256×256 mm^2^, flip angle = 8°, acquisition matrix size = 256×256.

### Image processing and calculation of cortical thickness

Cortical reconstruction was performed with the FreeSurfer software package (version 7.1.0) (https://surfer.nmr.mgh.harvard.edu). In brief, preprocessing included motion correction, intensity normalization, removal of non-brain tissue, Talairach transformation, segmentation of the subcortical white matter and gray matter volumetric structures, tessellation and smoothing of the white matter boundary, and automated topology correction and surface deformation following intensity gradients to optimally place the gray/white and gray/CSF borders defined at the location with the greatest shift in signal intensity (47-51). Cortical thickness was calculated based on the distance between white and gray matter boundaries at each vertex. The cortical surface was then parcellated into 68 regions based on the Desikan–Killiany (DK) atlas (52). All results of cortical parcellations were visually inspected, and inaccuracies in segmentation were manually edited. Histograms of all regions’ values for each site were also computed for visual inspection.

### Statistical analyses

Differences in demographics and clinical measurements between the chronic ketamine use group and control group were tested using t tests for continuous variables and chi-square tests of independence for categorical variables. Group cortical thickness differences for DK atlas regions between groups were examined using analysis of covariance (ANCOVA), with age, gender, educational level, smoking, drinking, and whole brain mean cortical thickness serving as covariates. To test whether cortical thickness differences in chronic ketamine users were potentially related to ketamine use characteristics, including age of ketamine use onset, the duration of ketamine use, and the quantity of ketamine consumption (estimated total lifetime ketamine consumption), Pearson correlations were used to assess associations of thickness of selected cortical regions with ketamine use profiles. All statistical analyses were performed with R version 4.0.3. Results were considered significant if the Bonferroni corrected p-value < 0.01.

## Results

### Sample Characteristics

**Table 1** presents demographics, addictive substance use (including ketamine use, cigarette smoking, alcohol drinking and other substance use) characteristics for patients with ketamine dependence/ketamine addicts/chronic ketamine users and drug-free healthy controls (HC). There were no significant group differences in age, gender, handedness, or marital status. However, ketamine addicts received fewer years of education than the control subjects. Smoking and drinking were more prevalent among the ketamine addicts. One-third of ketamine addicts also tried methamphetamine or other stimulants. There was no reported use of cocaine, heroin, or other drugs.

**Table 1.**
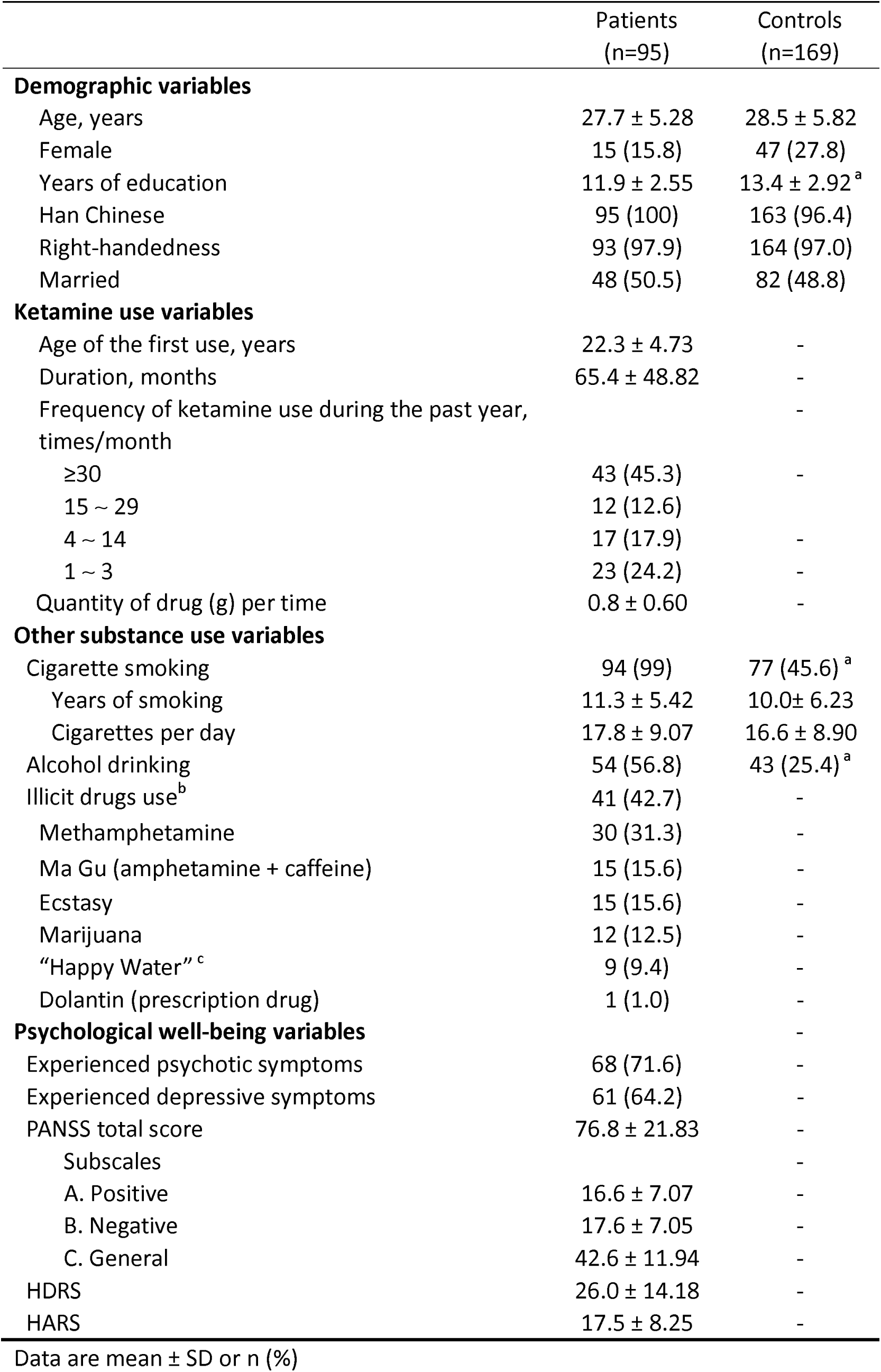
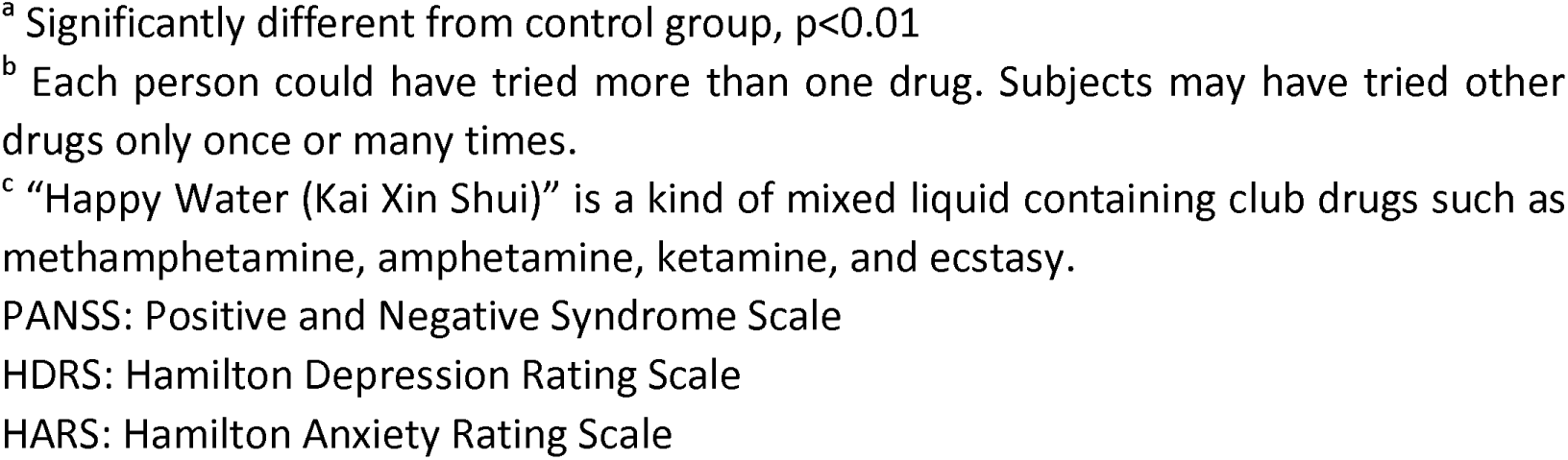
Demographic and clinical characteristics of patients with ketamine dependence and control subjects

|  | Patients<br>(n=95) | Controls<br>(n=169) |
| --- | --- | --- |
| **Demographic variables** |  |  |
| Age, years | 27.7 ± 5.28 | 28.5 ± 5.82 |
| Female | 15 (15.8) | 47 (27.8) |
| Years of education | 11.9 ± 2.55 | 13.4 ± 2.92<sup>a</sup> |
| Han Chinese | 95 (100) | 163 (96.4) |
| Right-handedness | 93 (97.9) | 164 (97.0) |
| Married | 48 (50.5) | 82 (48.8) |
| **Ketamine use variables** |  |  |
| Age of the first use, years | 22.3 ± 4.73 | - |
| Duration, months | 65.4 ± 48.82 | - |
| Frequency of ketamine use during the past year,<br>times/month |  | - |
| ≥30 | 43 (45.3) | - |
| 15 ~ 29 | 12 (12.6) |  |
| 4 ~ 14 | 17 (17.9) | - |
| 1 ~ 3 | 23 (24.2) | - |
| Quantity of drug (g) per time | 0.8 ± 0.60 | - |
| **Other substance use variables** |  |  |
| Cigarette smoking | 94 (99) | 77 (45.6)<sup>a</sup> |
| Years of smoking | 11.3 ± 5.42 | 10.0 ± 6.23 |
| Cigarettes per day | 17.8 ± 9.07 | 16.6 ± 8.90 |
| Alcohol drinking | 54 (56.8) | 43 (25.4)<sup>a</sup> |
| Illicit drugs use<sup>b</sup> | 41 (42.7) | - |
| Methamphetamine | 30 (31.3) | - |
| Ma Gu (amphetamine + caffeine) | 15 (15.6) | - |
| Ecstasy | 15 (15.6) | - |
| Marijuana | 12 (12.5) | - |
| “Happy Water”<sup>c</sup> | 9 (9.4) | - |
| Dolantin (prescription drug) | 1 (1.0) | - |
| **Psychological well-being variables** |  |  |
| Experienced psychotic symptoms | 68 (71.6) | - |
| Experienced depressive symptoms | 61 (64.2) | - |
| PANSS total score | 76.8 ± 21.83 | - |
| Subscales |  | - |
| A. Positive | 16.6 ± 7.07 | - |
| B. Negative | 17.6 ± 7.05 | - |
| C. General | 42.6 ± 11.94 | - |
| HDRS | 26.0 ± 14.18 | - |
| HARS | 17.5 ± 8.25 | - |

Data are mean ± SD or n (%)
<sup>a</sup> Significantly different from control group, $p < 0.01$
<sup>b</sup> Each person could have tried more than one drug. Subjects may have tried other drugs only once or many times.
<sup>c</sup> “Happy Water (Kai Xin Shui)” is a kind of mixed liquid containing club drugs such as methamphetamine, amphetamine, ketamine, and ecstasy.
PANSS: Positive and Negative Syndrome Scale
HDRS: Hamilton Depression Rating Scale
HARS: Hamilton Anxiety Rating Scale

The prevalence of psychotic symptoms was 71.6% (n = 68) in ketamine addicts. For positive symptoms, 35.8% (n = 34) had hallucinations (most of them experienced auditory verbal hallucinations and visual hallucinations), 18.9% (n = 18) had delusions (mainly persecutory delusions). Other commonly experienced symptoms including depressive symptoms (64.2%, n = 61), memory decline (47.4%, n = 45), and irritability (32.6%, n = 31).

### Cortex thickness reductions in patients with ketamine dependence

Compared with healthy control subjects, a total of 26 regions were found to show significant cortical thickness reductions in patients with ketamine dependence (Bonferroni corrected, p-value < 0.01). Increased cortical thickness was not observed in ketamine dependent patients. **Figure 1** shows significant cortical thinning regions in ketamine dependent patients. Regions are listed in **Table 2** in order of effect size, from the strongest to the weakest effect size. Regions with the most extensive cortical thickness reductions were located in the frontal (including the dorsolateral prefrontal cortex, DLPFC) and parietal (including the precuneus) lobes.

**Table 2.** Regional of significant cortical thinning in patients with ketamine dependence ranked by effect size

|  |  | Patients | Controls |  |  |  |
| --- | --- | --- | --- | --- | --- | --- |
| Hemisphere | Region | Cortical thickness (mm) |  | F | Effect size |  |
|  |  | Mean ± SD | Mean ± SD |  | Cohen's f | CI |
| Left | Superior parietal lobule | 2.13 ± 0.09 | 2.21 ± 0.09 | 41.50 | 0.40 | 0.30 - 0.51 |
| Right | Precuneus | 2.34 ± 0.11 | 2.42 ± 0.10 | 38.88 | 0.39 | 0.28 - 0.50 |
| Right | Superior parietal lobule | 2.14 ± 0.10 | 2.21 ± 0.09 | 38.20 | 0.39 | 0.28 - 0.49 |
| Right | Supramarginal gyrus | 2.46 ± 0.12 | 2.54 ± 0.11 | 36.64 | 0.38 | 0.27 - 0.48 |
| Right | Inferior parietal lobule | 2.39 ± 0.10 | 2.47 ± 0.10 | 36.37 | 0.38 | 0.27 - 0.48 |
| Left | Postcentral gyrus | 2.00 ± 0.09 | 2.07 ± 0.09 | 35.88 | 0.37 | 0.27 - 0.48 |
| Left | Precuneus | 2.34 ± 0.11 | 2.41 ± 0.10 | 30.63 | 0.35 | 0.24 - 0.45 |
| Right | Superior frontal gyrus | 2.72 ± 0.12 | 2.80 ± 0.11 | 30.06 | 0.34 | 0.24 - 0.45 |
| Left | Inferior parietal lobule | 2.36 ± 0.12 | 2.43 ± 0.11 | 26.60 | 0.32 | 0.22 - 0.43 |
| Left | Lateral occipital gyrus | 2.12 ± 0.10 | 2.18 ± 0.09 | 25.25 | 0.31 | 0.21 - 0.42 |
| Left | Superior frontal gyrus | 2.78 ± 0.11 | 2.85 ± 0.12 | 24.27 | 0.31 | 0.20 - 0.41 |
| Right | Rostral middle frontal gyrus | 2.29 ± 0.09 | 2.35 ± 0.10 | 23.48 | 0.30 | 0.20 - 0.41 |
| Right | Paracentral lobule | 2.48 ± 0.16 | 2.58 ± 0.15 | 22.99 | 0.30 | 0.19 - 0.40 |
| Right | Lateral occipital gyrus | 2.21 ± 0.09 | 2.26 ± 0.09 | 22.06 | 0.29 | 0.19 - 0.40 |
| Left | Supramarginal gyrus | 2.44 ± 0.13 | 2.51 ± 0.12 | 20.91 | 0.29 | 0.18 - 0.39 |
| Right | Inferior frontal gyrus (pars opercularis) | 2.56 ± 0.15 | 2.64 ± 0.14 | 20.80 | 0.29 | 0.18 - 0.39 |
| Left | Inferior temporal gyrus | 2.75 ± 0.14 | 2.82 ± 0.11 | 20.73 | 0.28 | 0.18 - 0.39 |
| Left | Fusiform gyrus | 2.66 ± 0.13 | 2.73 ± 0.09 | 20.57 | 0.28 | 0.18 - 0.39 |
| Left | Precentral gyrus | 2.57 ± 0.13 | 2.63 ± 0.12 | 17.64 | 0.26 | 0.16 - 0.37 |
| Right | Caudal middle frontal gyrus | 2.51 ± 0.12 | 2.58 ± 0.13 | 17.47 | 0.26 | 0.16 - 0.37 |
| Left | Inferior frontal gyrus (pars opercularis) | 2.52 ± 0.15 | 2.59 ± 0.13 | 17.23 | 0.26 | 0.15 - 0.36 |
| Left | Caudal middle frontal gyrus | 2.55 ± 0.14 | 2.62 ± 0.12 | 17.20 | 0.26 | 0.15 - 0.36 |
| Left | Rostral middle frontal gyrus | 2.30 ± 0.09 | 2.35 ± 0.09 | 16.64 | 0.25 | 0.15 - 0.36 |
| Left | Middle temporal gyrus | 2.76 ± 0.14 | 2.83 ± 0.13 | 16.60 | 0.25 | 0.15 - 0.36 |
| Left | Superior temporal gyrus | 2.72 ± 0.15 | 2.79 ± 0.14 | 16.40 | 0.25 | 0.15 - 0.36 |
| Right | Fusiform gyrus | $2.69 \pm 0.12$ | $2.74 \pm 0.11$ | 16.33 | 0.25 | 0.15 - 0.36 |

**Figure 1.**
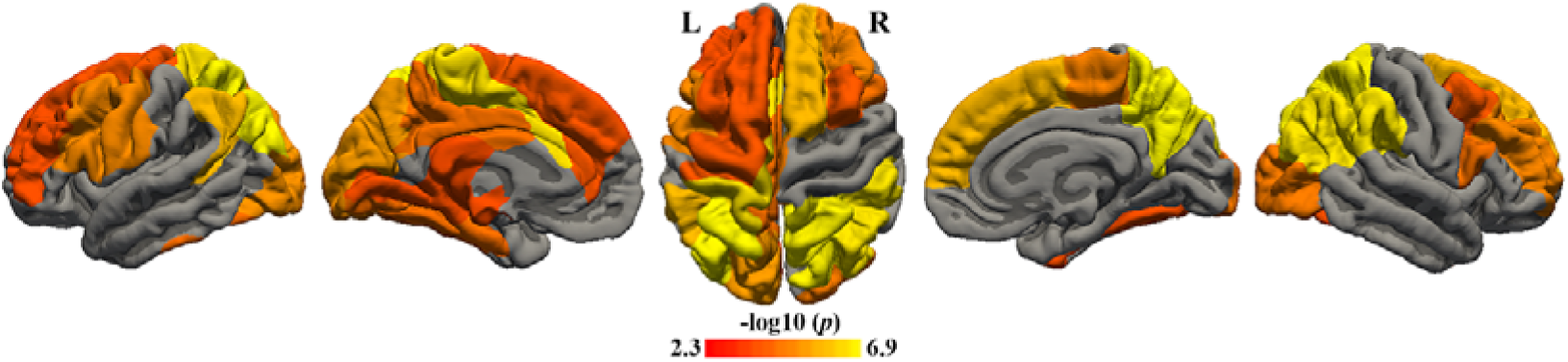
Cortical thickness reductions in patients with ketamine dependence

### Correlations between ketamine use variables and reduced cortical thickness regions

Pearson correlation was applied to assess the correlations between cortical thickness reductions in 26 regions and age of ketamine use onset, the duration of ketamine use (month), and the estimated total lifetime ketamine consumption (g). Correlation analysis found a negative correlation between estimated total lifetime ketamine consumption and cortical thickness in the right inferior parietal lobule (**Figure 2,** r = −0.41, corrected p < 0.001) and right rostral middle frontal gyrus (**Figure 3,** r = −0.38, corrected p = 0.004). However, this study failed to find correlations between age of ketamine use onset or the duration of non-medical ketamine use and regions of any significant thinning cortical thickness.

**Figure 2.**
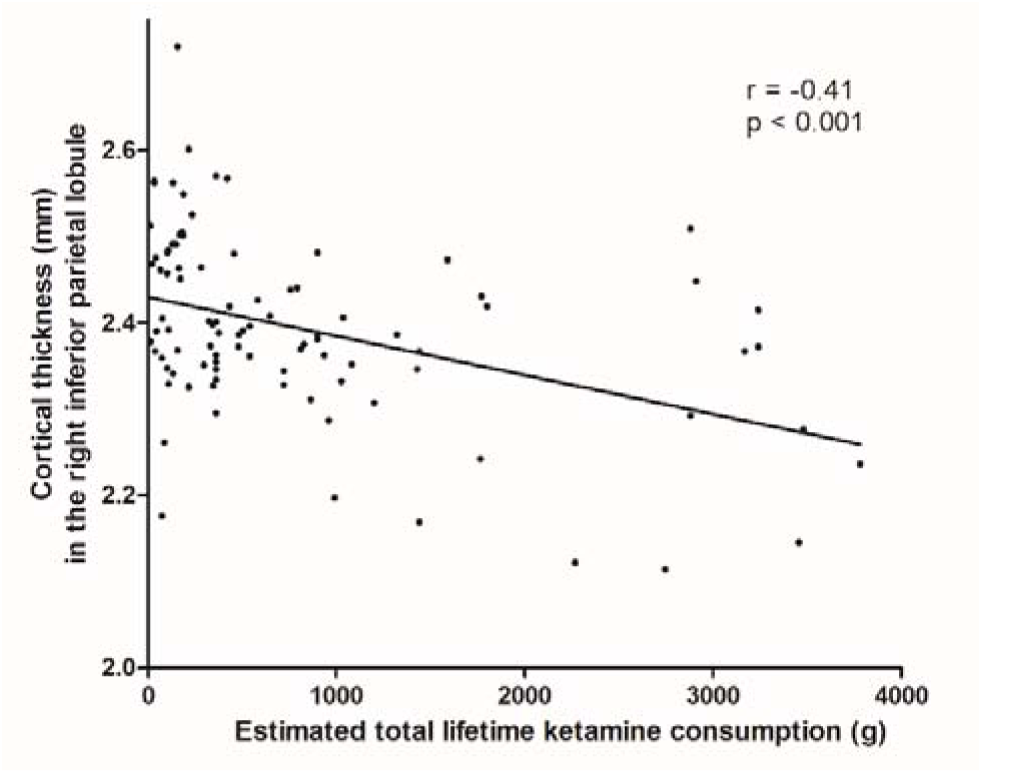
Corrections between total lifetime ketamine consumption and cortical thickness reductions of the right inferior parietal lobule in patients with ketamine dependence

**Figure 3.**
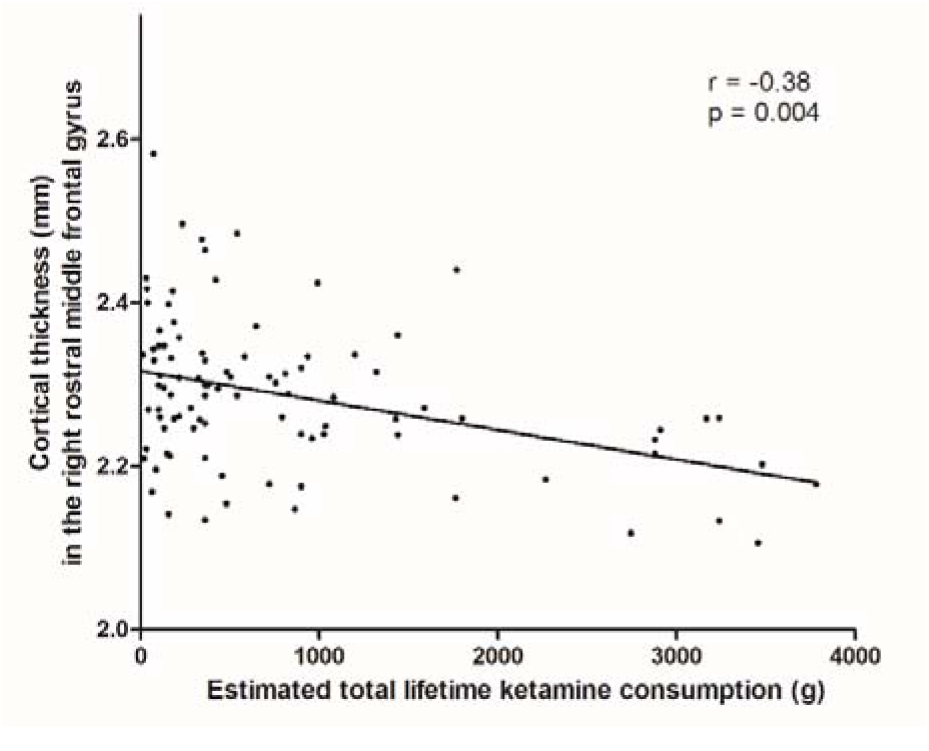
Association between total lifetime ketamine consumption and cortical thickness reductions of the right rostral middle frontal gyrus in patients with ketamine dependence

## Discussion

We sought to identify regions with cortical thickness alternations in chronic, non-medical ketamine users and to establish whether cortical thickness reductions were related to ketamine use characteristics. To our knowledge, this is the first study of cortical thickness abnormalities in patients with ketamine dependence/addiction. The main findings of this study are that patients with ketamine dependence/chronic ketamine users, compared with drug-free healthy volunteers, show the following: 1) impairments of mental well-being, including psychotic symptoms, and symptoms of depression; 2) widespread cortex thickness reductions, with the most extensive thickness reductions in the frontal (including the dorsolateral prefrontal cortex, DLPFC) and parietal (including the precuneus) lobes; 3) estimated total lifetime ketamine consumption (but not age of ketamine use onset, or the duration of non-medical ketamine use) is correlated with two specific cortical regions of cortical thickness reductions: the right inferior parietal lobule and the right rostral middle frontal gyrus.

### Impaired mental well-being in patients with ketamine dependence

The current study found that over 70% of patients with ketamine dependence had psychotic symptoms, including auditory verbal hallucinations, visual hallucinations, and persecutory delusions. And more than 60% of ketamine addicts experienced depressive symptoms. However, all the patients in this study were from rehabilitation centers, chronic use of ketamine-induced psychosis and the comorbidity of depression are more prevalent among treatment seeking ketamine addicts. Ketamine use related mental problems, such as hallucinations, agitation, and anxiety, were also reported among recreational ketamine users from the general population or sub-population (clubbers) (53). Furthermore, several clinical studies also indicates that long-term, non-medical ketamine use has been associated with poor psychological well-being, including psychotic symptoms and cognitive impairment (17, 54), out-of-body experiences (18), symptoms of depression and anxiety (19), sleeping problems (20). Preclinical studies found that repeated exposure to ketamine for a period of 14 days (55) or 28 (56) days can lead to cognitive impairments. The current study further indicates a link between long-term, non-medical use of ketamine and impaired mental well-being.

### Cortical thickness thinning in patients with ketamine dependence

This study found significant and consistent thinner cortices in a wide range of regions, with the most extensive thickness reductions in the frontal (including the dorsolateral prefrontal cortex, DLPFC) and parietal (including the precuneus) lobes, in patients with ketamine dependence. Thinning frontal cortical thickness reduction was also observed in individuals with other drug dependence, such as cocaine (57), heroin (58), and alcohol dependence (59), as well as non-drug /behavioral addiction, such as addiction to internet (60) and gaming (61). One interesting aspect of the present findings is that, compared with drug-free healthy controls, subjects with ketamine dependence only exhibited significantly decreased but no increased cortical thickness. However, both decreased and increased cortical thickness were observed in individuals with heroin dependence (58), as well as people with Internet gaming disorder (61) or internet addiction (60). These inconsistent findings of cortical thickness alternations may indicate that the consistent thinning cortical thickness might be a trait for ketamine-dependent patients.

The results of widespread reductions of cortical thickness in ketamine dependent subjects, with the most pronouncedly in the frontal and parietal areas (including prefrontal cortex and precuneus), are similar the findings of patients with schizophrenia who exhibit excessive cortical thinning in widespread areas of the brain, but most pronounced in the frontal and temporal areas (62, 63). The most extensive frontal cortical thinning in both patients with ketamine dependence and patients with schizophrenia may reflect the underline mechanisms of shared psychotic symptoms between chronic ketamine use and schizophrenia.

### Correlations between ketamine consumption and two reduced cortical thickness regions

This study found correlations between estimated total lifetime ketamine consumption and cortical thickness in the right inferior parietal lobule and the right rostral middle frontal gyrus. However, this study failed to find correlations between age of ketamine use onset or the duration of non-medical ketamine use and regions of any significant thinning cortical thickness. Our previous studies also showed correlations between estimated total lifetime ketamine consumption and alternations of gray matter (29) as well as white matter (30) in chronic ketamine users. The current findings suggest that heavier ketamine users may display stronger cortical thickness reductions in specific regions, which might be served as neuroimaging-based biomarkers for stratifying severity of ketamine use.

### Implications for ketamine’s long-term treatment for patients with depression

Ketamine has a long history of medical use for starting and maintaining anesthesia with safety and effectiveness. Ketamine, as a novel antidepressant, however, has some safety concerns. Although a small but growing body of highly consistent studies have demonstrated short-term use of ketamine’s unique, rapid, and robust antidepressant properties (64), even for older adults (aged 60 years or older) with RTD (65). Single and repeated ketamine infusions also decreased suicidal ideation in patients suffering from TRD (11, 66). Up to date, only a few studies have examined the safety of long-term use of ketamine in clinical setting. An open-label study has demonstrated the long-term (up to 1 year) safety and manageable tolerability of weekly or every-other-week dosing of Esketamine nasal spray (28-mg, 56-mg, or 84-mg) plus a new oral antidepressant for individuals with TRD (67). Furthermore, psychiatric, psychotomimetic and other side-effects were report after single and repeated doses of Esketamine in the treatment of depression (68). It remains unclear, if long-term administration of low-dose ketamine will increase propensity for ketamine addiction following antidepressant treatment. Also, there are still numerous unsolved problems in terms of its addictive properties and controversies regarding its role as antidepressant. Considering that the long-term safety remains largely unknown, Intranasal Esketamine for TRD is still controversial. For example, Esketamine for TRD is not recommended by The National Institute for Health and Care Excellence (NICE) in UK (69).

The current findings of cortical thickness reductions in patients with ketamine dependence provide evidence that long-term, medical use of ketamine should be cautious. It is important to balance the promise and risks of ketamine treatment for TRD in clinical setting (70). Further work is warranted to explore the exact mechanism behind ketamine’s ability to treat TRD, which may provide more valuable strategies to prevent ketamine’s potential addiction/abuse. On the other hand, future neurobiological studies that examine those thickness reduction regions and their functions, may help us to clarify the pathological mechanism for ketamine addiction and its medical properties.

### Limitations and future directions

Although this study has strength in terms of interpreting the findings, shows some similarity with the alternations of cortical thickness observed in schizophrenia, and sheds some light on the potential brain effects of long-term ketamine use for TRD, there are some potential limitations to this work that should be considered: 1) a notable limitation of this study was the retrospective cross-sectional design in the current study. It does not directly and unequivocally establish a causal relationship between ketamine use and alternations of cortical thickness reported. Whether cortical thickness abnormalities in ketamine addiction are only a reflection of chronic ketamine use, or also a pre-existing disposition to ketamine addiction? Whether other potential confounding influences, such as duration of abstinence and developmental timecourses from recreational use to dependence of ketamine, be associated with the cortical thickness reductions in individuals with ketamine dependence? Whether these deficits are reversible, and to what extent therapy or abstinence might reverse them? This cross-sectional design research provides little clearly insight into the above questions. Studies with a more comprehensive longitudinal design are needed to explore these confounding factors; 2) the two groups we studied were not matched for cigarette smoking, with a higher rate of cigarette smoking in ketamine-dependent subjects (although there were no significant group differences in years of smoking and smoked cigarettes per day among smokers). A line of study reported the reduced cortical thickness among cigarette smokers (71, 72). Thus, the analysis included smoking as a covariate. Also, we did not find significant association between years of smoking and cortical thickness reduction, and we found no correlation between any significant regions and estimated total smoked cigarettes, all p values > 0.1; 3), prevalence of alcohol drinking was also not well-matched between the two groups. Compared with healthy controls, both men and women with alcohol dependence showed reduced cortical thickness (59), especially in the prefrontal cortical region (73). However, this study excluded subject with alcohol dependence, and included drinking as a covariate in the analysis; 4) education levels were not well-matched. However, when we explicitly explored the impact of education level on cortical thickness reduction in the group of ketamine user, we found no statistically significant correlation (p > 0.1). Furthermore, we included it as a covariate in the analysis. These suggest that our findings cannot simply be explained in terms of this variable; 5) while none of the ketamine users in our sample had current clinical presentations or history of bipolar disorder, schizophrenia, or any other psychotic disorders, many had ketamine use-induced psychosis, and symptoms of depression and anxiety. Although there was a time link between ketamine use and the development of psychotic or depressive symptoms, we could not fully ascertain that these symptoms were caused by the use of ketamine; 6) confounding by other substances (especially stimulants) cannot be totally excluded, although, we did excluded subject with other drug dependence, except for nicotine; 7) as some ketamine dependent patients experienced severe psychotic symptoms or severe depressive symptoms, history of medications (such as antipsychotic medications and antidepressants) could not be totally excluded. However, in order to minimize medication effects and make patients more cooperative in the scanner, few ketamine users who had administration of medication within two weeks of scanning were included in this study.

Despite the harmful effects of non-medical use of ketamine, medical use of ketamine should never be discouraged or devalued. Compared with other antidepressants, ketamine is unique in its ability to rapidly help individuals suffering with severe TRD (74), especially with suicidal ideation (75). Evidence indicates that short-term medical use of ketamine appears to be safe in the short-term treatment of TRD (76). Furthermore, there is a wealth of evidence indicating the value of ketamine as an accessible and affordable medication for anesthesia (especially in less privileged medical facilities), and ketamine is also well-established for pain management, and critical care (5-7). Nevertheless, the findings of this study provide an important insight into the safety precautions of medical use of ketamine, especially when it is used in the long-term or with large amounts of quantity. Researchers should be warned that there are many ketamine use related issues that need more investigation, especially matters related to addictive potential and strategies to mitigate illicit use, and adverse effects of brain damage of long-term administration.

### Conclusion

In conclusion, the findings from this study provide evidence that long-term, non-medical use of ketamine linked to cortical thickness reductions, especially ketamine addicts with great total lifetime ketamine consumption. This study provides a probe to advance an understanding of its addiction and provides insights in the safety of repeated ketamine infusions for the treatment of depression. A greater understanding of the underlying mechanism of cortical thickness reductions in chronic ketamine users/ketamine addicts could lead to the markable improvement of risk/benefit ratios of Esketamine for individuals with TRD.

## Data Availability

Data will be available by request.

## Acknowledgments and Disclosures

This study was supported by the National Natural Science Foundation of China (grant no. 81871057 to JT, 81671324 to TL and 81671325 to YL), and the Hunan Provincial Natural Science Foundation of China (grant no. 2020JJ4794 to YL and 2020JJ4795 to TL), and by the “Hundred Talents Program” funding from Zhejiang University. The funders had no role in study design, data collection and analysis, decision to write the report or to submit the paper for publication.

We thank all subjects who participated in this study.

The authors declare no biomedical financial interests or potential conflicts of interest.

